# Anorexia Nervosa Register-based Severity Trajectories: Polygenic and Comorbid Associations

**DOI:** 10.64898/2026.09.28.26364163

**Authors:** Mohamed Abdulkadir, Janne Tidselbak Larsen, Zi-Ping Zhang, Katherine L. Musliner, Loa Clausen, Cynthia M. Bulik, Liselotte Vogdrup Petersen, Zeynep Yilmaz

## Abstract

**Importance:** Anorexia nervosa (AN) is a severe psychiatric disorder with heterogeneous outcomes and elevated mortality. Characterizing severity trajectories may improve early risk stratification and targeted intervention.

**Objective:** To identify AN severity trajectories using a register-based severity index (AN-RSI) and examine their associations with polygenic scores (PGS), preceding (prior to AN diagnosis) as well as subsequent (beyond 5 years post AN diagnosis) psychiatric and somatic disorders.

**Design:** Population-based cohort study.

**Setting:** Nationwide study in Denmark.

**Participants:** Individuals born between 1963 and 2007 with diagnosed AN (N = 9534 individuals). Genetic data were available in a subsample (N = 5126).

**Exposures:** Forty-one PGS indexing traits relevant to AN, and psychiatric and somatic disorders prior to AN diagnosis.

**Main Outcomes and Measures:** AN-RSI trajectories, psychiatric, and somatic disorders beyond 5 years post AN diagnosis.

**Results:** Three trajectories were identified: “low and stable” (32.6%), “medium and stable” (53.5%), and “high increasing” (14.0%). The “low and stable” group was used as the comparison group for all analyses. Each 1 standard deviation (SD) increase in AN PGS corresponded with 13% greater odds of following the “high increasing” trajectory. Increases in BMI and attention-deficit hyperactivity disorder (ADHD) PGS corresponded with 15% and 14% lower odds of following the “high increasing” trajectory, respectively. No preceding psychiatric or somatic disorders were significantly associated with trajectory membership. In contrast, subsequent psychiatric and somatic disorders were more likely in individuals following the “high increasing” trajectory with odds ratios ranging between 1.7 (ADHD) and 3.7 (psychoactive substance dependence) for psychiatric disorders and between 1.6 (poor fetal growth of child) and 4 (microcytic anemia) for somatic disorders.

**Conclusions and Relevance:** Our findings demonstrate that individuals with AN follow distinct severity trajectories that are associated with differences in polygenic risk and post-diagnostic morbidity patterns but not with pre-diagnosis psychiatric or somatic morbidity. As the field progresses, incorporating PGS into comprehensive assessments at first presentation may aid in early identification, personalized intervention, and resource allocation for individuals at higher risk of developing a severe illness trajectory.

**Key points:** *Question:* Do patients with anorexia nervosa follow distinct severity trajectories that are associated with specific genetic and morbidity risks?

*Findings:* In this register-based study we identified three severity trajectories of anorexia nervosa. Those who followed the most severe illness trajectory had greater genetic liability to lower body weight and anorexia nervosa and were at greater risk for developing subsequent psychiatric and somatic disorders.

*Meaning:* Illness severity trajectories in anorexia nervosa marked by genetic risk may assist in early identification and intervention for those at greatest risk of severe illness.

## Introduction

Anorexia nervosa (AN) is a severe psychiatric disorder associated with substantial morbidity, mortality, and societal burden^1,2^. Despite treatment advances, long-term prognosis remains poor, with many patients experiencing chronic illness trajectories, relapse, and severe medical and psychological complications ^3–6^.

One of the key challenges in improving outcomes for AN lies in the heterogeneity of its course and severity. The literature clearly shows an association between severe AN and elevated mortality risk ^7,8^. Patients exhibit wide variation in symptom progression, treatment response, and long-term outcomes, making it difficult to identify those at greatest risk for severe and persistent illness. Previous studies often relied on cross-sectional assessments or short-term follow-up, which may fail to capture the dynamic nature of the disorder over time. Consequently, there is a need for longitudinal data on disease progression, highlighting the value of a data-driven method that can more accurately characterize AN progression and identify meaningful trajectories based on illness severity ^9^.

AN is a heritable disorder, with twin studies estimating heritability between 0.28 and 0.74 ^10^. This broad range may reflect differing inclusion criteria, with narrower criteria yielding more homogeneous groups with fewer psychiatric comorbidities and higher heritability estimates ^11^. Genetic studies of AN have predominantly focused on case status, ^12–16^ with only one study investigating the genetic correlates of AN severity ^16^. This partly reflects the scarcity of severity data in samples powered for genetic analyses.

To address this gap, we recently developed a register-based severity index for AN (AN-RSI)^7^. This index follows individuals in national health registers from their first AN diagnosis; using it, we have shown that individuals with severe AN have higher medication prescription rates and mortality risk than those with less severe AN, indicating that the AN-RSI captures clinically relevant aspects of the illness ^7,17^.

Integrating comprehensive longitudinal AN-RSI data with genetic data offers a unique opportunity to move toward a more nuanced understanding of the genetic correlates of disease trajectories. Longitudinal investigations can be enriched by incorporating polygenic scores (PGS)—a single continuous score obtained from genome-wide association studies (GWAS) that indexes the genetic liability of an individual to a particular trait ^18^. Relating several PGS that index different traits (e.g., depression, anxiety, insulin sensitivity, body mass index [BMI]) with severity trajectories enables investigation of unique genetic profiles associated with severity trajectories and may allow for early detection of those who are more likely to follow a severe illness course.

To this end, the present study used the AN-RSI framework to construct severity trajectories during the first five years following an initial AN diagnosis. This initial period is likely to shape long-term outcomes and may reveal distinct patterns of illness development. Specifically, the study: (1) identifies distinct AN-RSI trajectories; (2) examines associations between these trajectories and predisposing risk factors, including PGS of psychiatric and somatic disorders; and (3) determines subsequent emergence of psychiatric and somatic disorders associated with different trajectories. By elucidating these trajectories, we aim to enhance early risk stratification, inform personalized interventions, and ultimately improve long-term outcomes for individuals affected by AN.

## Methods

### Study population

We included individuals born between 1 January 1963 and 31 December 2007 who received an International Classification of Diseases 8^th^ revision (ICD-8; 306.50) or 10^th^ revision (ICD-10; F50.0, F50.1) diagnosis of AN after their 6^th^ birthday ^7^. We excluded individuals who emigrated or died within five years of diagnosis or who had less than five years of follow-up before study end on 31 December 2018 (see Figure S1 for study flow-chart). The genetic analyses (N = 5126) comprised a subsample of AN cases (born after 1981) from the Anorexia Nervosa Genetics Initiative-Denmark (ANGI-DK) and the Eating Disorders Genetics Initiative-Denmark (EDGI-DK) ^19,20^.

### Severity index

The AN-RSI is described in detail elsewhere ^7^. Briefly, it is a composite of six register-derived indicators: early onset, late onset, inpatient admissions, outpatient treatment episodes, treatment length, and illness duration, weighted in collaboration with experienced clinicians ^7,17^.

### Construction of trajectories

We constructed AN-RSI trajectories using latent class analysis with the FlexMix package in R ^21^. Yearly AN-RSI scores for the first five years after diagnosis were modeled with a Poisson distribution, given their count nature. Models included birth year and sex as covariates and a quadratic time term to account for nonlinearity. Solutions with up to five classes were compared using the Bayesian Information Criterion, Akaike Information Criterion, entropy, class size (avoiding classes <5% of the sample), and interpretability.

### Selection of phenotypes for association analyses

All available ICD-8 and ICD-10 codes were included; ICD-8 codes were converted to ICD-10 using a unidirectional mapping ^22^, and ICD-10 codes were mapped to phecodes using phecodeX ^23^. Diagnoses during the five-year trajectory period were not evaluated. Somatic and psychiatric phecodes occurring prior to AN diagnosis were considered exposures, whereas those occurring after the trajectory period were considered outcomes.

### Genetic data

#### Genotyping

Dried blood spots from routine screening of nearly all Danish newborns since 1981 are stored at the Danish Newborn Screening Biobank ^24^. Genotyping has been described elsewhere ^25^. This study was approved by the Danish Data Protection Agency, the Danish Scientific Ethics Committee, the Danish Health Data Authority, and the Danish Newborn Screening Biobank Steering Committee. The Danish Scientific Ethics Committee waived informed consent in accordance with Danish legislation on biobank-based research.

#### Polygenic scores

We selected 41 PGS (Table S1) previously associated with AN ^3,13,14^, covering psychiatric (n = 11; including obsessive-compulsive disorder [OCD], schizophrenia, major depressive disorder [MDD]), anthropometric (n = 13; including BMI, fat mass, and fat-free mass), metabolic (n = 12; including insulin sensitivity index, type 2 diabetes mellitus, and high cholesterol), and educational (n = 2; educational attainment and cognitive performance) traits as well as PGS indexing physical activity time, multisite chronic pain, and age at menarche. PGS were calculated using LDpred2 (36 traits) or meta-polygenic risk scores (i.e., attention-deficit hyperactivity disorder [ADHD], autism spectrum disorder [ASD], bipolar disorder, MDD, and schizophrenia) ^26,27^. Calculation of PGS was restricted to individuals of European ancestry as previously described ^28^.

### Statistical Analyses

Associations between PGS and AN-RSI trajectories were tested using multinomial regression adjusted for sex and the first five ancestry-informative principal components (see Figure S2 for analytical design of study). Phecodes occurring before AN diagnosis were analyzed using multinomial regression and those occurring after the trajectory period using logistic regression, both adjusted for birth year and year of AN diagnosis. Phecode analyses were stratified by sex because some phecodes are sex-specific (e.g., irregular menstrual cycle); sex-neutral analyses included both sexes after removing sex-specific phecodes. To ensure robustness, we excluded phecodes with fewer than 15 cases in any trajectory group. Consequently, male-only analyses were not possible given the small number of males; males were, however, included in the sex-neutral analyses. We corrected for multiple hypothesis testing using the Benjamini-Hochberg false discovery rate (FDR) method ^29^. Significance was defined as FDR-adjusted Q < 0.05.

## Results

### Sample description

The study population included 9534 individuals (93.55% female) with a mean age at first AN diagnosis of 19.37 years (standard deviation [SD]: 6.28; Table 1). The median (interquartile range [IQR]) AN-RSI was 1 (IQR = 2) in year 1, 2 (IQR = 2) in year 2, 2 (IQR = 4) in year 3, 2 (IQR = 5) in year 4, and 3 (IQR = 5) in year 5.

**Table 1:** Descriptives of sample following first anorexia nervosa (AN) diagnosis.

|  | N | % of total | Posterior probability<sup>1</sup> mean | Female % | Age at diagnosis mean (SD) | First year AN-RSI median (IQR)<sup>2</sup> | Second year AN-RSI median (IQR)<sup>2</sup> | Third year AN-RSI median (IQR)<sup>2</sup> | Fourth year AN-RSI median (IQR) | Fifth year AN-RSI median (IQR)<sup>2</sup> |
|---|---|---|---|---|---|---|---|---|---|---|
| Total sample | 9534 | 100 | - | 93.6 | 19.4 (6.3) | 1(2) | 2(2) | 2(4) | 2(5) | 3(5) |
| Low and stable trajectory | 3107 | 32.6 | 0.98 | 93.7 | 19.3 (5.6) | 0 (1) | 0 (1) | 0 (1) | 0 (1) | 0 (1) |
| Medium and stable trajectory | 5096 | 53.5 | 0.98 | 93.2 | 19.7 (6.7) | 2 (1) | 2 (1) | 3 (2) | 3 (3) | 4 (4) |
| High increasing trajectory | 1331 | 13.9 | 0.97 | 94.7 | 18.4 (6.0) | 3 (4) | 8 (4) | 10 (6) | 13 (8) | 14 (9) |

SD: standard deviation. RSI: register-based severity index. IQR: interquartile range.
<sup>1</sup>The mean probability of the latent class model accurately predicting class membership for individuals in that class.
<sup>2</sup>Year of illness following first diagnosis.

### AN-RSI trajectories

A three-class model fit the data best, with better fit statistics than lower-order solutions and adequately sized classes compared with four- and five-class solutions (Table S2). Most cases were assigned to the “medium and stable” trajectory (N = 5096, 53.45%; see Table 1 and Figure 1) followed by the “low and stable” trajectory (N = 3107, 32.59%) and the “high increasing” trajectory (N = 1331, 13.96%). Median yearly AN-RSI scores were 0 for the “low and stable” trajectory, 2–4 for the “medium and stable” trajectory, and 3–14 for the “high increasing” trajectory. The “low and stable” trajectory was used as the referent for all analyses.

**Figure 1:**
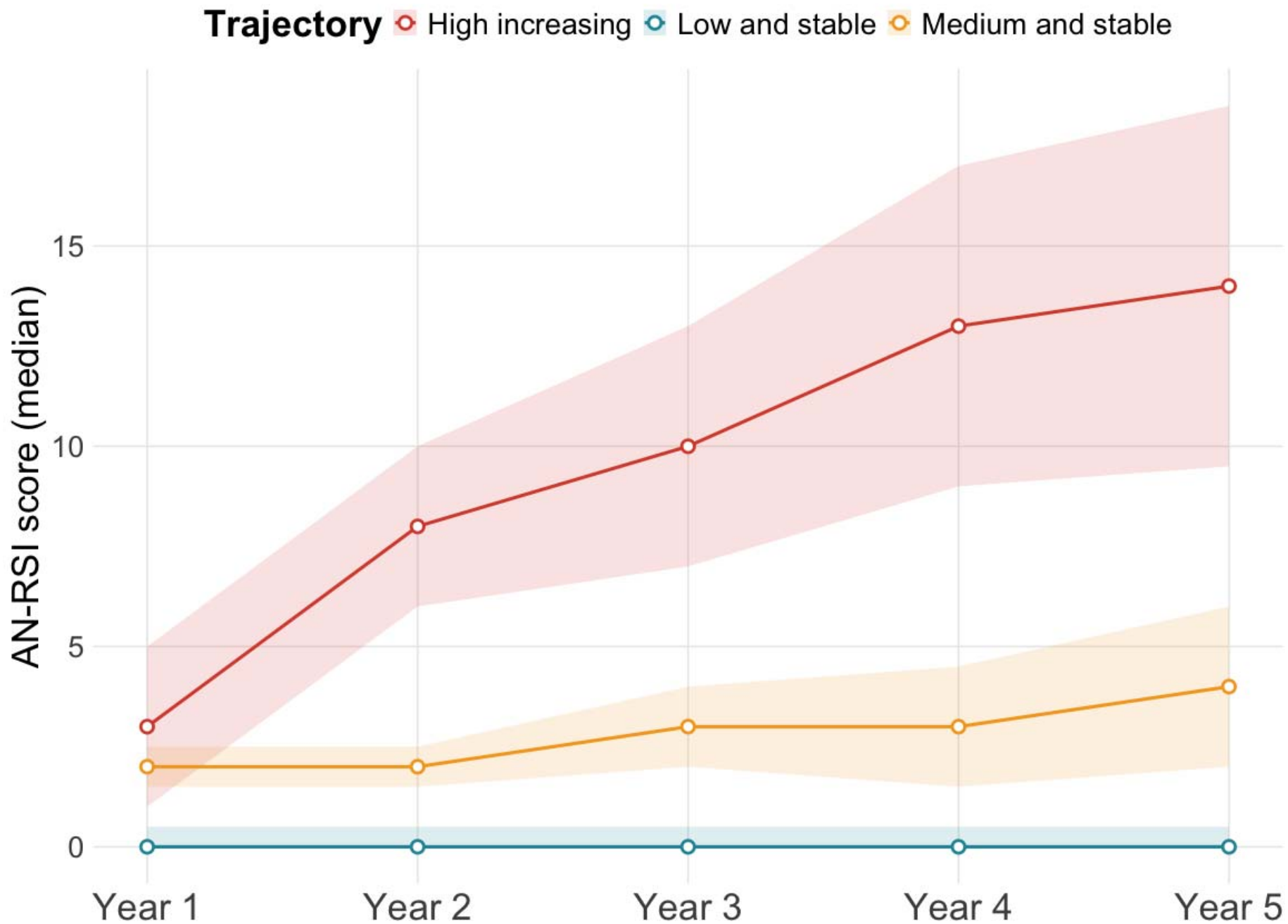
Trajectories of anorexia nervosa (AN) register-based severity index (RSI) in the first five years following AN diagnosis. Solid lines depict the median AN-RSI for the “high increasing” trajectory (N =1331) in red, the “medium and stable” trajectory (N = 5096) in orange, and the low and stable trajectory (N = 3107) in green. The shaded areas represent the interquartile ranges.

### Association of PGS with AN-RSI trajectories

Of the 5126 individuals in the PGS analyses, 2507, 1880, and 739 followed the “medium and stable”, “low and stable”, and “high increasing” trajectories, respectively (for descriptives see Table S3).

Each 1-SD increase in BMI, fat mass, fat-free mass, and weight PGS was associated with 15% to 35% decreased odds of following the “high increasing” trajectory (Figure 2; see Table S4 for full results). Higher AN PGS corresponded with 13% increased odds of following the “high increasing” trajectory. In contrast, higher ADHD PGS corresponded with 14% decreased odds of following the “high increasing” trajectory.

**Figure 2:**
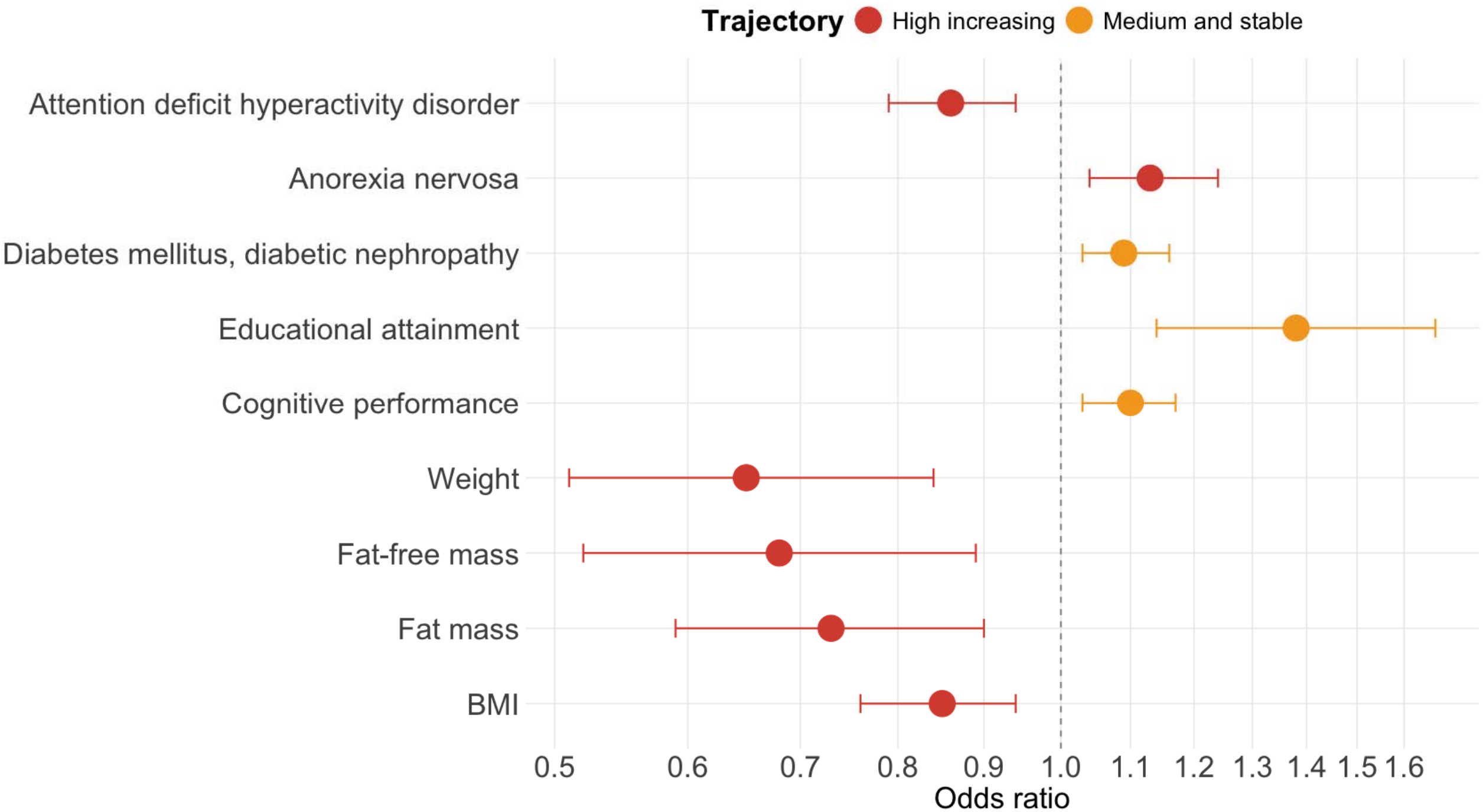
Associations between polygenic scores (PGS) and anorexia nervosa (AN) register-based severity index (RSI) trajectories. The “medium and stable” trajectory (N = 2507) is depicted in orange and the “high increasing” trajectory (N = 739) in red; the “low and stable” trajectory (N = 1880) was used as referent. Depicted are the associations that remained significant after correcting for multiple hypothesis testing (false discovery rate adjusted Q < 0.05). Associations are depicted as odds ratios per 1 standard deviation increase in the PGS with dots representing the odds ratio point estimate and the l ne depicting the 95% confidence interval.

**Figure 3:**
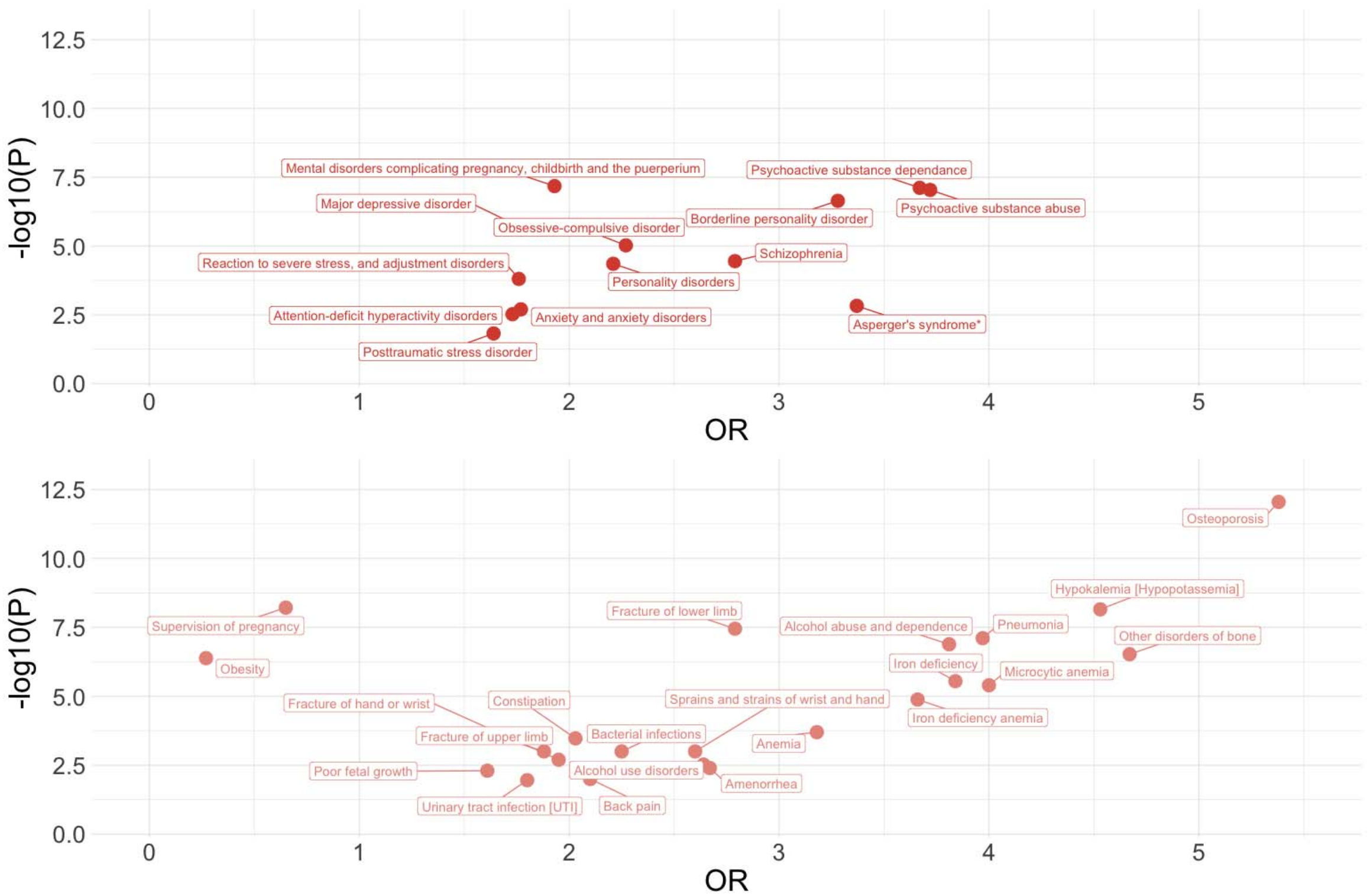
Results from the association analyses with anorexia nervosa register-based severity index “high increasing” trajectory as exposure and subsequent (five years after first anorexia nervosa diagnosis) psychiatric (top panel) and somatic (bottom panel) phecodes as outcome in females. The "low and stable" trajectory was used as referent. Depicted are the -log10 p-values for each association between the “high increasing” trajectory (N = 1261) and a phecode with at least 15 cases. Only associations that are significant after correcting for multiple hypothesis testing (false discovery rate adjusted Q < 0.05) are shown (see Table S7 for full results). OR = odds ratio.

**Figure 4:**
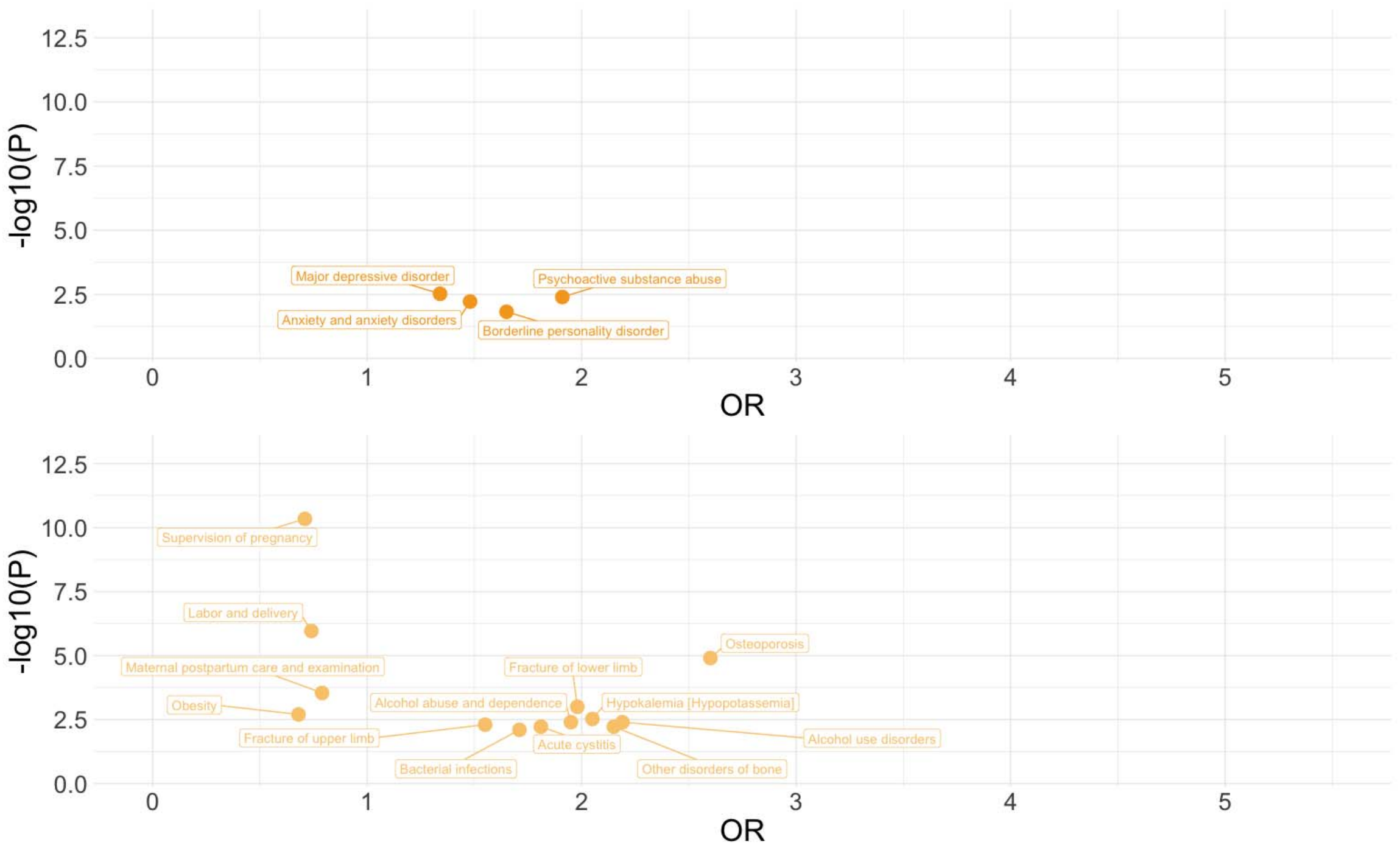
Results from the association analyses with anorexia nervosa register-based severity index “medium and stable” trajectory as exposure and subsequent (five years after first anorexia nervosa diagnosis) psychiatric (top panel) and somatic (bottom panel) phecodes as outcome in females. The "low and stable" trajectory was used as referent. Depicted are the -log10 p-values for each association between the “high increasing” trajectory (N = 4748) and a phecode with at least 15 cases. Only associations that are significant after correcting for multiple hypothesis testing (false discovery rate adjusted Q < 0.05) are shown (see Table S7 for full results). OR = odds ratio.

PGS associated with increased odds of the “medium and stable” trajectory were educational attainment (38%), cognitive performance (10%), and diabetic nephropathy (9%).

### Phecodes prior to AN-RSI trajectories

After filtering, 21 and 20 phecodes were analyzed in the female-only and sex-neutral analyses, respectively (Tables S5-S6). None of the included phecodes were significantly associated with AN-RSI trajectories.

### Phecodes after the AN-RSI trajectories

A total of 87 and 66 phecodes met the filtering threshold for the female-only and sex-neutral analyses, respectively (Tables S7-S8). In the female-only analysis, 37 phecodes were significantly (Q < 0.05) associated with the trajectories (Table S7); 21 of them uniquely associated with the “high increasing” trajectory, 3 of them uniquely associated with the “medium and stable” trajectory and the remaining 13 associated with both trajectories. Patterns were similar in the sex-neutral analyses (Table S8); we therefore report female-only results.

The “high increasing” trajectory—but not the “medium and stable” trajectory—showed increased risk for several psychiatric disorders including schizophrenia, borderline personality disorder, Asperger’s syndrome (merged into ASD in ICD-11), OCD, and ADHD; odds ratios (ORs) ranged from 1.7 (ADHD) to 3.7 (psychoactive substance dependence). Several other psychiatric disorders (e.g., alcohol abuse and dependence, psychoactive substance abuse, and anxiety disorders) were associated with both trajectories with ORs ranging from 1.3 (MDD) to 3.8 (alcohol abuse and dependence).

Risk for several somatic disorders—such as pneumonia, iron deficiency, amenorrhea, back pain, and constipation—was increased only in individuals who followed the “high increasing” trajectory with ORs ranging from 1.6 (poor fetal growth of child) to 4.0 (microcytic anemia). Risk for other somatic disorders (e.g., bacterial infections, hypokalemia, other disorders of the bone) was increased in both the “high increasing” and “medium and stable” trajectories with ORs ranging from 1.6 (fracture of upper limb) to 5.4 (osteoporosis). We also observed lower odds for supervision of pregnancy in both trajectories (“high increasing” OR = 0.6; “medium and stable” OR = 0.7). Furthermore, the “high increasing” and the “medium and stable” trajectories had 73% and 32% lower odds of receiving an obesity diagnosis, respectively.

## Discussion

In this nationwide Danish population study, we identified three AN severity trajectories that are significantly and differentially associated with genetic factors and subsequent psychiatric and somatic disorders. The “high increasing” trajectory carried a significant polygenic profile highlighted by lower anthropometric (BMI, fat mass, fat-free mass, weight), lower ADHD, and higher AN PGS compared to the “low and stable” trajectory. We investigated both predisposing phecodes and subsequent phecodes of the three trajectories and found associations only with psychiatric and somatic morbidity following, not preceding, AN-RSI trajectories, emphasizing the need for continued clinical monitoring, especially for those following a “high increasing” trajectory.

BMI is a complex trait with behavioral and metabolic genetic components, with signals enriched in the central nervous system and linked to energy metabolism ^30,31^. Our findings extend the consistently reported negative genetic correlations between AN and anthropometric traits to AN severity ^12–14^. This contrasts with a smaller Swedish register-based study in which higher BMI PGS was associated with greater AN severity ^16^. This discrepancy likely reflects different severity definitions: the Swedish study used the self-reported Clinical Impairment Assessment, which measures psychosocial impairment due to eating disorder symptoms ^16^. The AN-RSI instead mainly reflects treatment seeking and duration.

To our knowledge, this is the first study to report a positive significant association between AN PGS and AN severity. The associations of both BMI and AN PGS with the most severe trajectory are clinically relevant because both PGS are associated with poor childhood growth in the general population ^32^. Therefore, poor childhood growth observed together with high AN PGS and low BMI PGS could serve as early indicators of AN severity risk.

Higher ADHD PGS was associated with lower odds of following the most severe trajectory. The relationship between AN and ADHD is unclear, with most studies reporting positive but others negative genetic correlations ^14,33–36^. The inverse association may reflect perfectionism, which is typical of AN but not of ADHD ^37–39^. Alternatively, higher ADHD PGS may be linked to fewer hospital contacts; in another Danish register study of early-onset MDD, higher ADHD PGS predicted brief rather than prolonged or persistent hospital contact ^40^.

Higher educational attainment PGS was associated only with the “medium and stable” trajectory after correction for multiple comparisons. A longstanding discussion exists regarding the association between educational attainment and AN ^41–43^. The genetic correlation between AN and educational attainment is estimated at 0.25 to 0.27 ^13,14^. Epidemiological studies also suggest a positive association ^37,44–46^. One Mendelian randomization analysis showed a bidirectional positive relationship between AN and higher educational attainment ^47^. This was likely due to horizontal pleiotropy, whereby variants associated with higher educational attainment also increase AN liability ^47,48^. This shared genetic basis may reflect traits such as perfectionism and need for order and organization—both of which are advantageous for higher educational attainment ^49,50^.

Individuals who followed the most severe illness course were at greatest risk for morbidity after the trajectory period. Overall, phecodes that showed significant association with the AN-RSI trajectories broadly reflect two categories: psychiatric disorders and somatic disorders that are likely attributable to weight loss and malnutrition (e.g., osteoporosis, anemia, iron deficiency, hypokalemia, and amenorrhea) ^8^. Phecodes in both categories were more frequent in the “high increasing” trajectory, further illustrating within-diagnosis heterogeneity.

These results extend our previous finding that the AN-RSI is associated with subsequent psychiatric disorders at the ICD-10 block level ^7^. Using a phecode approach, we show that the AN-RSI trajectories are associated with specific psychiatric disorders, including OCD, Asperger’s syndrome, substance dependence, and borderline personality disorder for the “high increasing” trajectory, and MDD and anxiety for both the “high increasing” and the “medium and stable” trajectories. This concurs with evidence that psychiatric comorbidity is the rule rather than the exception in AN ^2,14,35,51,52^; in our sample, it was most pronounced in the “high increasing” trajectory.

The positive phenotypic association between the “high increasing” trajectory and ADHD contrasts with our ADHD PGS finding. One explanation is that phenotypic analyses only captured ADHD diagnosed after the trajectory period; the mean age at ADHD diagnosis in this sample was 32 years, well above the typical age (around 12 years) ^53^. Adult-onset ADHD may differ genetically and environmentally from childhood-onset ADHD ^36,54,55^. Alternatively, ADHD may be masked in childhood, as it is diagnosed less often in girls than boys but at comparable rates in adult men and women ^56^. Increasing AN severity may make masked ADHD clinically apparent in women following the most severe trajectory.

### Strengths and limitations

This study has several strengths, including the large sample size and the availability of detailed longitudinal health data from Danish national registers alongside genotype data. Our study investigated both psychiatric and somatic disorders alongside the AN-RSI trajectories, giving a more comprehensive overview of morbidity in AN patients. Furthermore, Danish healthcare is free of charge, making our findings less susceptible to selection bias. We included males in our sex-neutral analyses to improve generalizability to both sexes. Our findings should be interpreted in the context of several limitations. Our sample did not include enough males to obtain robust estimates from male-only analyses. We were unable to separately evaluate the restricting and binge/purge subtypes of AN as the register data did not contain information on these subtypes. We also did not evaluate the impact of comorbid conditions that were diagnosed during the initial 5 years following the first AN diagnosis (the period in which the trajectories were established); these diagnoses likely shaped the trajectories but were omitted from our analyses to establish a clear temporal association between the psychiatric and somatic diagnoses and the AN-RSI trajectories. Individuals in more severe trajectories had more hospital contacts, which may have increased the likelihood of other conditions being diagnosed (detection bias). Excluding individuals who died or emigrated within five years of diagnosis may also have removed some of the most severe cases. Furthermore, our genetic study was limited to individuals of European ancestry, which could impact the generalizability of our findings to other ancestries. Finally, although the AN-RSI is a novel measure allowing register-based research on severity, its scores were based on hospital diagnoses, and the weighting of the items that comprised the total score was based on clinical judgment and may not replicate perfectly in different clinical settings or patient populations.

## Conclusion

Our findings are potentially clinically relevant, demonstrating that individuals with AN follow distinct severity trajectories that have different genetic risk as measured by PGS. Crucially, increased genetic liability for AN and for lower body weight may represent an early indicator of severe AN illness trajectory risk. Currently, we have no tools to reliably identify the likely AN course at first presentation—brief remitting illness, medium duration and severity, or severe and enduring. The clinical utility of PGS is gaining attention and, although the effect sizes observed here are modest, PGS may in future complement initial assessments and aid early identification and treatment planning for those at highest risk of severe illness ^57^. Further research and more robust PGS based on larger AN sample sizes will aid in building such prediction models to personalize treatment and improve AN treatment outcomes.

## Author contributions

Dr. Abdulkadir had full access to all of the data in the study and takes responsibility for the integrity of the data and the accuracy of the data analysis. **Concept and design:** Yilmaz and Petersen**. Acquisition, analysis, or interpretation of data:** all authors. **Drafting of the manuscript:** Abdulkadir, Petersen, Yilmaz. **Critical review of the manuscript for important intellectual content:** all authors. **Statistical analyses:** Abdulkadir. **Obtained funding**: Yilmaz, Petersen, Bulik. **Supervision:** Yilmaz, Petersen.

## Data Sharing Statement

Summary statistics that were used in deriving the polygenic scores are available through the GWAS catalog (refer to the PubMed IDs in Table S1) or as weights on figshare (https://doi.org/10.6084/m9.figshare.23597019.v1). The Danish registry data used in this study are under restricted access and protected by Danish legislation and can therefore not be shared. The genetic data from the ANGI-DK and EDGI-DK are also under restricted access and protected by Danish legislation; access can only be granted after approval by the iPSYCH Data Access Committee on the secured Danish computing cluster GenomeDK (https://genome.au.dk/).

