## Supplementary material for "Anorexia Nervosa Register-based Severity Trajectories: Polygenic and Comorbid Associations": Figure S1

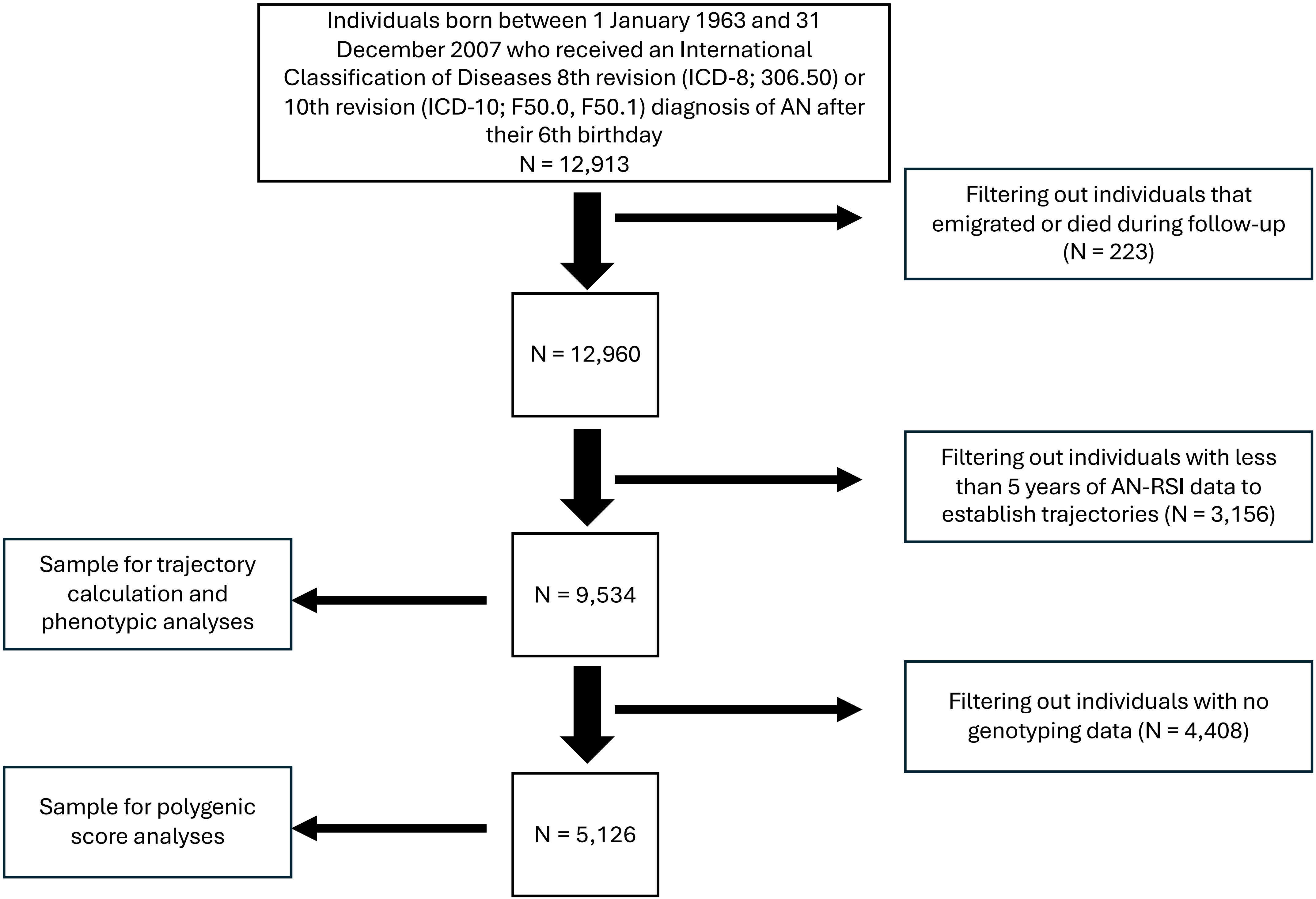
Individuals born between 1 January 1963 and 31 December 2007 who received an International Classification of Diseases 8th revision (ICD-8; 306.50) or 10th revision (ICD-10; F50.0, F50.1) diagnosis of AN after their 6th birthday
N = 12,913
Filtering out individuals that emigrated or died during follow-up (N = 223)
N = 12,960
Filtering out individuals with less than 5 years of AN-RSI data to establish trajectories (N = 3,156)
Sample for trajectory calculation and phenotypic analyses
N = 9,534
Filtering out individuals with no genotyping data (N = 4,408)
Sample for polygenic score analyses
N = 5,126
