## Supplementary material for "Anorexia Nervosa Register-based Severity Trajectories: Polygenic and Comorbid Associations": Figure S2

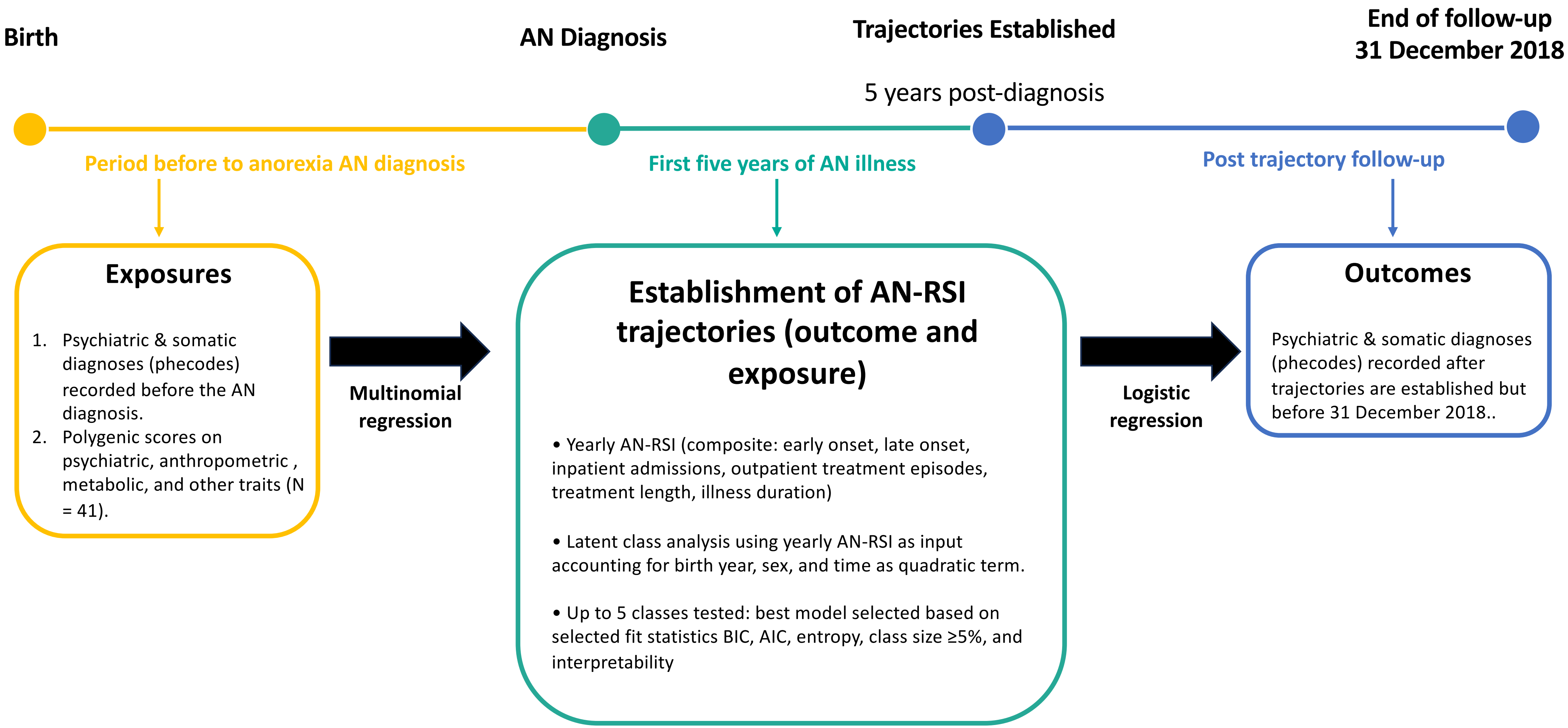
Birth
AN Diagnosis
Trajectories Established
5 years post-diagnosis
End of follow-up
31 December 2018
Period before to anorexia AN diagnosis
First five years of AN illness
Post trajectory follow-up
Exposures
1. Psychiatric & somatic diagnoses (phecodes) recorded before the AN diagnosis.
2. Polygenic scores on psychiatric, anthropometric , metabolic, and other traits (N = 41).
Multinomial regression
Establishment of AN-RSI trajectories (outcome and exposure)
• Yearly AN-RSI (composite: early onset, late onset, inpatient admissions, outpatient treatment episodes, treatment length, illness duration)
• Latent class analysis using yearly AN-RSI as input accounting for birth year, sex, and time as quadratic term.
• Up to 5 classes tested: best model selected based on selected fit statistics BIC, AIC, entropy, class size ≥5%, and interpretability
Logistic regression
Outcomes
Psychiatric & somatic diagnoses (phecodes) recorded after trajectories are established but before 31 December 2018..
